# DISRUPTIONS IN LIVELIHOODS, HEALTHCARE ACCESS AND HEALTH OUTCOMES IN WEST AFRICA DURING EMERGING INFECTIOUS DISEASE OUTBREAKS

**DOI:** 10.1101/2024.04.27.24306478

**Authors:** Rashid Ansumana, Alfred S. Bockarie, Monya Konneh, Najmul Haider, Eniola Cadmus, Alberta Korvah, Joyce Fayiah, Yolaine Kate Waka-Metzger, Angella M. George, Gassimu Mallah, Joseph M. Lamin, Osman Dar, Susannah Mayhew, Doris Bah, Pepe Bilivigoi, Nfaly Maggasouba, Simeon Cadmus, Robyn Alders

**Affiliations:** School of Public Health, College of Medical Sciences, Njala University, Bo Campus; Institute of Environmental Management and Quality Control, School of Environmental Sciences, Njala University, Njala Campus; Keele University, United Kingdom; University of Gamel Abdel Nasser of Conakry, Conakry Guinea; National Public Health Institute of Liberia, Monrovia, Liberia; University of Ibadan, Ibadan, Nigeria; London School of Hygiene and Tropical Medicine, UK; Australia National University; Ministry of Health, Guinea; Ministry of Health, Sierra Leone

**Author notes:** **Correspondence:** Rashid Ansumana(PhD), School of Public Health, College of Medical Sciences, Njala University.

## Abstract

**Background:** Outbreaks of infectious diseases like Ebola virus disease, Lassa fever, and COVID-19 have severely strained infrastructural systems and social services across West Africa. We investigated the disruptions caused by emerging outbreaks on access to healthcare, health outcomes, and livelihoods in West Africa.

**Method:** A mixed-methods approach was utilized, conducting extensive studies in Nigeria, Sierra Leone, Guinea, and Liberia through structured questionnaires, in-depth interviews with key informants, and focus group discussions. Using a device-to-cloud system guided by GIS for randomized sampling across the four nations, this technique allowed us to comprehensively analyze implications across imposed lifestyle changes on health and wellbeing due to disrupted healthcare.

**Results:** Our findings indicate drastic shifts in food consumption patterns and healthcare access. In Guinea, self-reported “Once Daily” meals astonishingly surged from 148 to 775 individuals (p<0.001), with analogous substantial increases observed in Liberia and Sierra Leone. Nigeria exhibited a varied response, with notable rises both in “Once Daily” and “Twice Daily” meal frequencies (p<0.001), reflecting broad dietary adaptations out of necessity. Additionally, there was a significant decrease in consumption of traditional protein sources like bushmeat, beef, and mutton, mainly because of disrupted supply chains and heightened concerns over food insecurity. Conversely, fish consumption slightly fell possibly due to its perceived safety or accessibility amidst the outbreak.

Healthcare services faced severe disruptions, particularly acute in Sierra Leone and Liberia compared to Guinea and Nigeria. The interruption of services drastically impacted everything from immunization rates to mental health, with a rise in reported anxiety and depression alongside public dissatisfaction towards the healthcare disruptions.

**Conclusion:** This study demonstrates the dramatic effects of infectious disease outbreaks on health access and diets in West Africa. The research calls for integrating health initiatives with social protection to strengthen the resilience of societies to meet all types of health challenges. It is imperative districts establish robust health systems and social security mechanisms to counter future public health crises.

## BACKGROUND

The intricate relationship between health and socio-economic well-being is deeply rooted in the fabric of human society. At the confluence of these critical dimensions lies the healthcare system, a fundamental pillar that ensures a population’s health and vitality and acts as a sentinel for broader socio-economic stability. West Africa, with its unique socio-cultural, economic, and political landscapes, has unfortunately been a recurrent epicentre for infectious disease outbreaks, notably the Ebola Virus Disease (EVD) in 2014-2016(Ansumana et al., 2014; Ansumana et al., 2017), Lassa fever (Ogbu, Ajuluchukwu and Uneke, 2007; Tambo, Adetunde and Olalubi, 2018; Kofman, Choi and Rollin, 2019) and, more recently, the COVID-19 pandemic(Ansumana, Sankoh and Zumla, 2020; Sumner, Hoy and Ortiz-Juarez, 2020). These outbreaks, while primarily health crises have invariably cast ripple effects, perturbing healthcare access, undermining livelihoods, and precipitating adverse health outcomes.

The importance of consistent and robust healthcare access cannot be overstated. In the context of West Africa, healthcare access is not only pivotal for managing endemic diseases such as malaria, tuberculosis, and HIV/AIDS but also for providing essential maternal and child health services(Smith and Whittaker, 2014). Outbreaks tend to exacerbate challenges faced by healthcare systems in resource-poor countries.

The economic backbone of many West African nations is agriculture, which employs a significant portion of the population. During outbreaks, lockdowns, travel restrictions, and market closures disrupt agricultural activities and supply chains. This has far-reaching consequences, including job loss, decreased income, and food insecurity, especially in rural areas(Egeru et al., 2020; Kipchumba et al., 2023).

For instance, during the EVD outbreak, many healthcare facilities in affected countries either closed or significantly reduced their operations due to fear of transmission, lack of protective equipment, or workforce attrition(Parpia *et al*., 2016). This curtailment reduced access to essential health services, increasing morbidity and mortality from non-Ebola causes(J W T Elston *et al*., 2017).

Furthermore, the socio-economic ramifications of outbreaks in West Africa are profound. Livelihoods, particularly in informal sectors, are often the first to be affected. As movement restrictions and lockdowns are implemented to curb the spread of disease, daily wage earners, small-scale traders, and artisans face immense challenges in sustaining their income(Vinck *et al*., 2019). This economic disruption, in turn, affects purchasing power and exacerbates food insecurity, further compromising vulnerable populations’ nutritional and health status(WFP, 2020).

Animal health is linked to human health and livelihoods, particularly in regions like West Africa. The concept of ‘One Health’ recognises that the health of people, animals, and the environment is interconnected(Conrad, Meek and Dumit, 2013). Outbreaks often have zoonotic origins, where infectious diseases are transmitted between animals and humans. For instance, the EVD outbreak is believed to have originated from bats and potentially transmitted to humans via the consumption of bushmeat(Olival and Hayman, 2014). Similarly, other pathogens with potential for outbreaks, such as Lassa and Marburg infectious disease and monkeypox, have animal reservoirs(Mylne *et al*., 2015). Disruptions caused by outbreaks can impact veterinary care and animal vaccination programs, elevating the risk of disease transmission between animals and humans.

Additionally, many of West Africa’s livelihoods depend on livestock and agriculture. Disease outbreaks among livestock can thus compound the socioeconomic challenges posed by human health crises(Rich and Wanyoike, 2010). Recognizing and addressing the intersection of animal and human health is essential for a comprehensive understanding of outbreak impacts and for devising effective intervention strategies.

Moreover, the broader health outcomes associated with these disruptions are multifaceted. Beyond the immediate morbidity and mortality from the outbreak, there is a cascade of secondary health implications. Reduced immunization coverage, for example, predisposes populations to vaccine-preventable diseases(Cooper *et al*., 2023). Additionally, mental health issues, often overlooked, emerge as significant concerns, with people grappling with anxiety, depression, and post-traumatic stress exacerbated by stigma and societal disruption(Shultz *et al*., 2016).

This paper aims to elucidate the interconnections and implications of disruptions in healthcare access, livelihoods, and health outcomes in West Africa during outbreaks. Through a comprehensive review of the literature and primary data, we will explore the multi-dimensional impacts of these disruptions and reflect upon strategies to mitigate their consequences.

## METHODS

### Study Design

This study adopted a mixed-methods approach, blending quantitative and qualitative data collection techniques, to delve into the disruptions in healthcare access, livelihoods, and health outcomes in West Africa amid outbreaks. Both prospective and retrospective data gathering were undertaken, offering a holistic depiction of the healthcare landscape during the outbreak episodes.

### Setting and Population

The research spanned four pivotal West African countries: Nigeria, Sierra Leone, Guinea, and Liberia. These countries were deliberately chosen considering their distinct experiences with outbreaks, enabling a multifaceted perspective on healthcare disruptions.

### Data Collection Tools and Strategy

**A** structured survey was curated and disseminated via the Open Data Kit (ODK) platform for the quantitative dimension. Responses from these surveys were securely stored and managed on the ONA server at https://ona.io/waohecostudy.

Parallelly, the qualitative dimension encompassed in-depth interviews and focus group discussions designed to extract richer insights into individual and community experiences, perceptions, and recommendations surrounding healthcare disruptions.

Random sampling was employed to ensure representativeness across diverse population subsets. The survey was directed using Geographic Information Systems (GIS), where geolocations of potential participants were randomly chosen. This GIS-directed approach enabled a spatially representative sample, ensuring that diverse geographical areas and their associated populations were adequately represented.

### Sample Size Determination

Using the OpenEpi tool, each country’s sample sizes were independently deduced, anchored on their respective SARS-CoV-2 seroprevalence rates. These computations were designed to achieve a 95% confidence interval (CI), factoring in elements like design effect, projected response rates, and possible contingencies such as non-responses or partial data.

### Quality Assurance

To maintain the highest data quality and ensure methodological rigor, daily dashboard monitoring was instituted to track and review incoming data in real time.

Surveyors documented their activities with photographs taken on-site, serving as a validation mechanism. Regular supervisory site visits were conducted to oversee the data collection process. Peer monitoring was employed, where teams cross-checked and validated each other’s work, ensuring consistency and accuracy.

### Data Analysis

Quantitative datasets were subjected to rigorous statistical analysis, with descriptive statistics elucidating the overall trends and inferential statistics employed for hypothesis testing and inference derivation.

Concurrently, qualitative data, sourced from interviews and discussions was transcribed verbatim. Thematic content analysis was then performed to discern emergent patterns and themes contextualized within the overarching healthcare disruption narrative.

### Ethical Considerations

Before their involvement, all participants rendered informed consent. Utmost confidentiality was observed throughout, with data being anonymised or pseudonymised. Ethical clearance was sought and obtained from relevant ethical review boards across the participating nations.

## RESULTS

### Outbreaks and Meal Frequency

We assessed the impact of an outbreak (SARS-CoV-2) on meal frequency patterns in Guinea, Liberia, Sierra Leone, and Nigeria. We categorized meal frequencies as ‘Once Daily,’ ‘Twice Daily,’ and ‘Thrice Daily,’ conducting statistical analyses to determine the significance of the changes observed (Table 1).

**Table 1:** Meal Frequency Table.

| Country | Meal Frequency | Before Outbreak | During Outbreak | P-Value |
| --- | --- | --- | --- | --- |
| Guinea | Once Daily | 148 | 775 | 0.00 |
|  | Thrice Daily | 3687 | 3078 | 0.00 |
|  | Twice Daily | 1721 | 1689 | 0.72 |
| **Liberia** | Once Daily | 2178 | 4345 | 0.00 |
|  | Thrice Daily | 310 | 70 | 0.00 |
|  | Twice Daily | 3222 | 1280 | 0.00 |
| **Nigeria** | Once Daily | 11 | 98 | 0.00 |
|  | Thrice Daily | 1212 | 755 | 0.00 |
|  | Twice Daily | 244 | 634 | 0.00 |
| **Sierra Leone** | Once Daily | 1806 | 2872 | 0.00 |
|  | Thrice Daily | 523 | 203 | 0.00 |
|  | Twice Daily | 3374 | 2620 | 0.00 |

In Guinea, there was a statistically significant increase in individuals consuming ‘Once Daily’ meals, from 148 before the outbreak to 775 during the outbreak (p < 0.001). Liberia displayed a similar trend in the increase of ‘Once Daily’ meal frequency, from 2,178 to 4,345 occurrences (p < 0.001). Sierra Leone followed this regional pattern, with ‘Once Daily’ meal frequency rising from 1,806 to 2,872 (p < 0.001). A significant reduction was also noted in ‘Twice Daily’ meals, from 3,374 to 2,620 (p < 0.001).

Nigeria, while experiencing an increase in ‘Once Daily’ meal frequency from 11 to 98 (p < 0.001), showed an increase in ‘Twice Daily’ meals from 244 to 634 (p < 0.001).

The observed increase in ‘Once Daily’ meal frequencies across all countries examined suggests a region-wide shift in dietary patterns due to the outbreak’s impact, echoing the findings of global health crises in the literature(Hendriks *et al*., 2022; Koyratty *et al*., 2022, 2022; Marti and Puertas, 2022; Lewis *et al*., 2023). The significant reduction in ‘Thrice Daily’ meals is consistent with patterns observed in food-scarce situations elsewhere in the world(González-Monroy *et al*., 2021).

The decline in ‘Twice Daily’ meals in Liberia, Sierra Leone and Guinea, contrary to the Nigerian data, indicates varied national responses to the outbreak. The reduction may reflect a more severe disruption of food systems compared to Nigeria, where an increase in ‘Twice Daily’ meals could suggest a different form of dietary adaptation, perhaps due to varying economic resilience or policy responses(Belton *et al*., 2021; Schreiber *et al*., 2022).

Statistically significant p-values affirm that these dietary changes are associated with the outbreak conditions, supporting the theory that health crises exert considerable influence on food security and nutritional behaviors (Beyene, 2023). These changes underscore the necessity for robust food security interventions during health emergencies to prevent long-term adverse health outcomes.

### Outbreaks and Household Protein Source Consumption in West Africa

Our investigation into household protein source consumption across Sierra Leone, Liberia, Nigeria, and Guinea provides critical insights into the dietary impacts of the recent outbreak. Consistent with disruptions observed in food systems during public health emergencies, there was a notable decline in the consumption of various protein sources during the outbreak(Béné, 2020; FAO, 2021).

**Fig 1:**
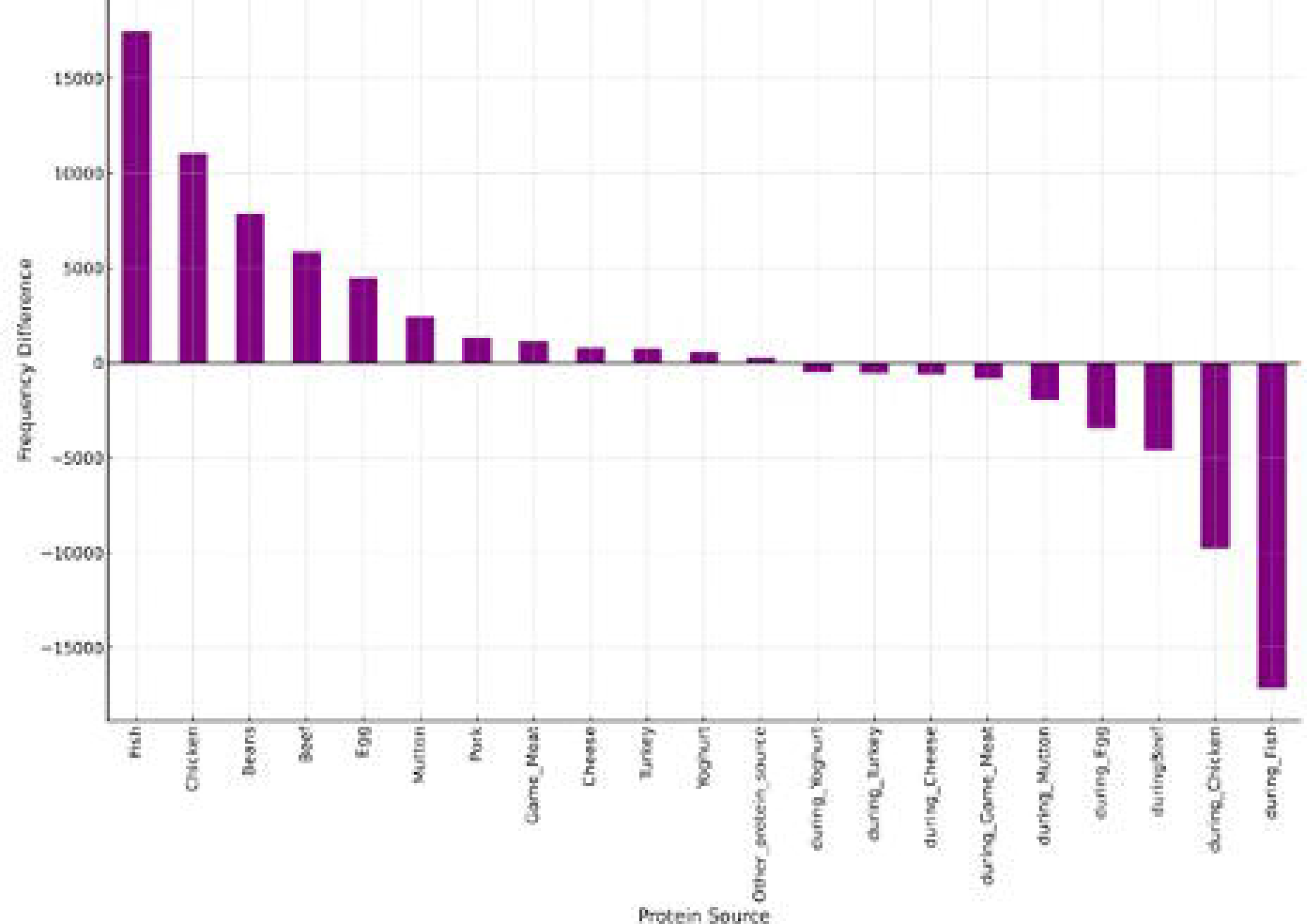
Change in Protein Source Consumption During SARS-CoV-2.

Before the outbreak, our data showed that common protein sources included bushmeat, beef, and mutton(Fig.1). In the subsequent period, the intake of these proteins significantly decreased, with the most pronounced reduction noted for bushmeat. This aligns with previous studies that have highlighted bushmeat as a potential zoonotic risk, particularly in West African contexts (Karesh et al., 2005; Wolfe et al., 2005). The decline in consumption of beef and mutton may be attributed to disrupted supply chains and heightened food security concerns, phenomena which are well-documented during health crises(World Bank, 2020).

Interestingly, the data suggest a more moderate decrease in the consumption of fish, which might indicate a strategic pivot towards protein sources perceived to be safer or more readily available during the outbreak(Headey and Martin, 2016). The differential changes across the various protein sources suggest a complex interplay of factors influencing household dietary choices during the outbreak(Barrett, 2010).

The observed shifts in consumption patterns underscored the profound effect of health crises on food security and dietary habits, emphasizing the need for robust food systems that can adapt to such challenges and protect vulnerable populations in the West African region(FAO, 2018).

Further, our qualitative study from West Africa suggests a complex interplay between community skepticism towards health interventions and the practical aspects of food safety and handling, with implications for the spread of foodborne diseases. Direct quotes from the study encapsulate the concerns: In Sierra Leone, it was noted that “Community beliefs about disease transmission from wild animals could lead to changes in hunting practices and dietary habits, affecting traditional food sources.” The study further records that lockdowns brought significant challenges: “During lockdowns, community members find it difficult to feed their animals due to restrictions on movement, potentially impacting livestock health and food availability.” Similarly, the situation in Nigeria mirrored these difficulties with animals also subjected to movement restrictions by their owners. The repercussions in Liberia included shifts in social behaviors: “Altered social norms and practices during outbreaks, such as reduced communal gatherings, can impact food distribution and consumption patterns within communities.” In addition, the spread of misinformation about the origins of diseases from certain foods has resulted in altered dietary choices: “Misinformation or beliefs about disease origins from certain foods could lead to changes in dietary habits, affecting demand for specific food items.” These insights from the field reveal the breadth and depth of the pandemic’s impact on traditional ways of life and the potential risks that arise from altered food-related practices.

### Healthcare Service Interruptions in West African Countries

During the SARS-CoV-2 pandemic, healthcare service interruptions were observed across several countries. We focused our analysis on Sierra Leone, Liberia, Guinea, and Nigeria to understand the extent of these interruptions.

In Sierra Leone, Liberia, and Guinea, the patterns of healthcare service interruptions were notably similar (Fig 2). The three most frequently interrupted services in these countries were: immunization, malaria treatment and routine bed net distribution.

Despite exhibiting a similar trend with immunization being the most interrupted service, Nigeria displayed distinct differences in the ranking of other services compared to Sierra Leone, Liberia, and Guinea.

The chart below provides a visual representation of the healthcare service interruptions in these countries:

**Fig. 2:**
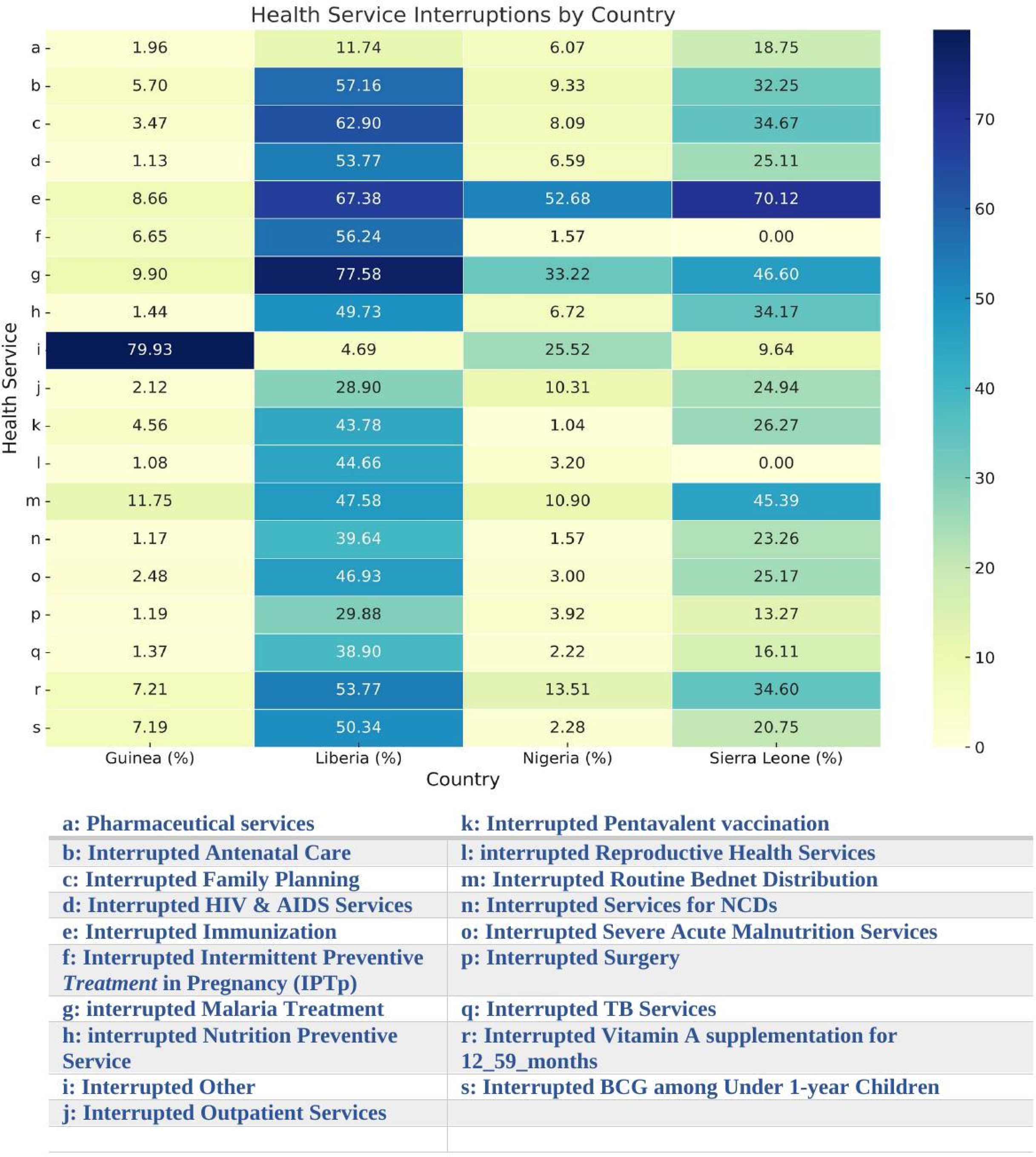
Healthcare Services Disruption Reported by Country.

By quantifying the interruptions, our analysis reveals the significant impact of the SARS-CoV-2 pandemic on essential healthcare services in West Africa. The disruption of crucial services, such as immunisation, underscores the cascading effects of global health crises on regional healthcare systems.

Liberia and Sierra Leone had more disruptions than Guinea and Nigeria (Fig 3).

**Fig. 3:**
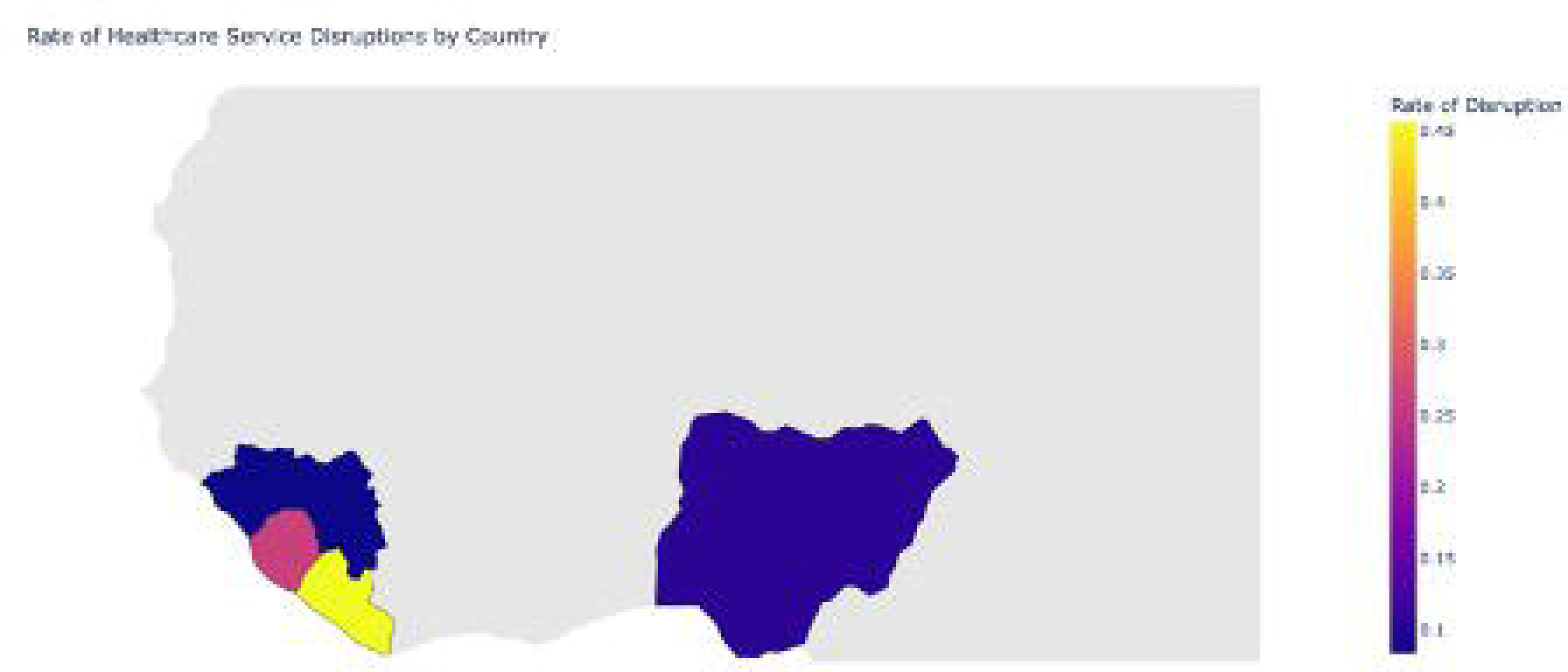
Choropleth Map of Healthcare Service Disruptions in West Africa.

Additional in-depth analyses of one country, Sierra Leone, which had a high level of disruptions revealed further the extent of service disruptions by outbreaks such as SARS-CoV-2.

### Disruptions in HIV Health Services

Our detailed examination of the repercussions of the SARS-CoV-2 pandemic on specific HIV services between 2019 and 2020 underscores significant variations in service provision (Fig 4).

**Fig 4:**
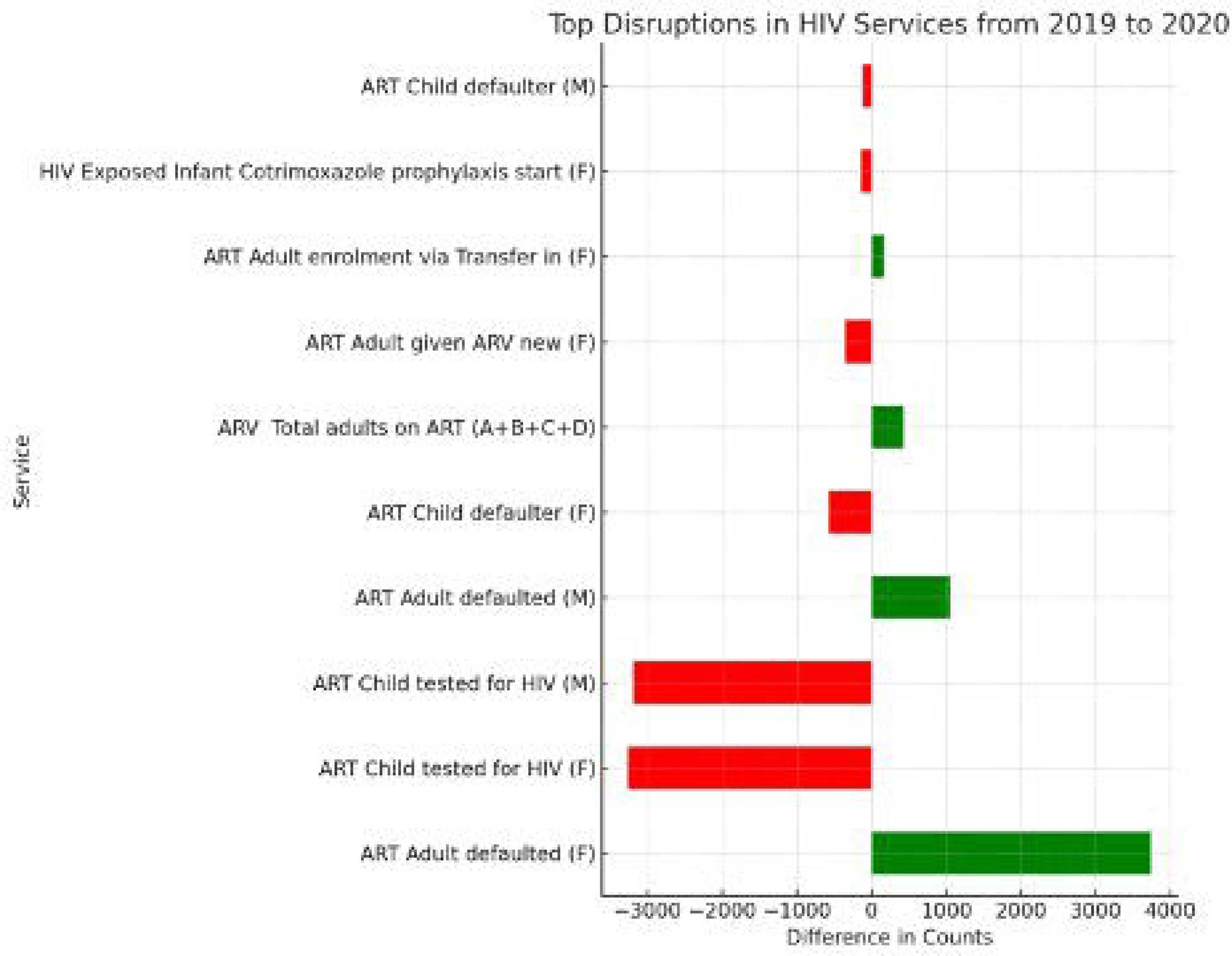
Disruptions in HIV Services.

The care and services provided to HIV-exposed infants were noticeably impacted: There was a pronounced decline observed in the “HIV-exposed infant follow-up visit” service, indicating potential barriers to continuous care during the early phase of the pandemic. The “HIV-exposed infant first visit” also faced a substantial reduction, suggesting delays or challenges in initial care access for this vulnerable group.

For adult HIV treatment, while some adult services highlighted resilience, others indicated vulnerabilities: “ART Adult defaulted” for both males and females increased, emphasizing potential challenges in maintaining treatment adherence during the pandemic. Interestingly, “ART Adult given ARV new” for both genders witnessed growth. This could be interpreted as an increase in new HIV diagnoses or improved access to treatment during this challenging period. Child HIV Treatment services demonstrated mixed outcomes: The “ART Child defaulter” service showcased stability, with only a slight decline, indicating the relative robustness of treatment adherence mechanisms for children.

On the contrary, “Child HIV positive Cotrimoxazole prophylaxis start” experienced a decrease, suggesting potential disruptions in preventive care for HIV-positive children.

In treatment transfers on treatment changes, the “ART Adult switched to 2nd line” service, a marker for treatment change, showed a significant drop. This could point to potential barriers to accessing alternative treatments when required.

Moreover, while the SARS-CoV-2 pandemic introduced challenges across the board in HIV services during 2020, the degree and nature of these disruptions varied widely. Specific services, especially those related to infants and treatment changes, faced pronounced challenges. These findings emphasize the importance of bolstering resilience in healthcare systems, especially for vulnerable demographics, in the face of global crises.

### Disruptions in TB Services

We observed a decline in both the number of HIV-positive TB patients receiving antiretroviral therapy (ART) and those receiving co-Trimoxazole preventive therapy from 2019 to 2020. This trend is likely indicative of the disruptions caused by the COVID-19 pandemic, which affected healthcare services globally, leading to challenges in accessing treatment for TB patients (Fig.5)

**Fig. 5:**
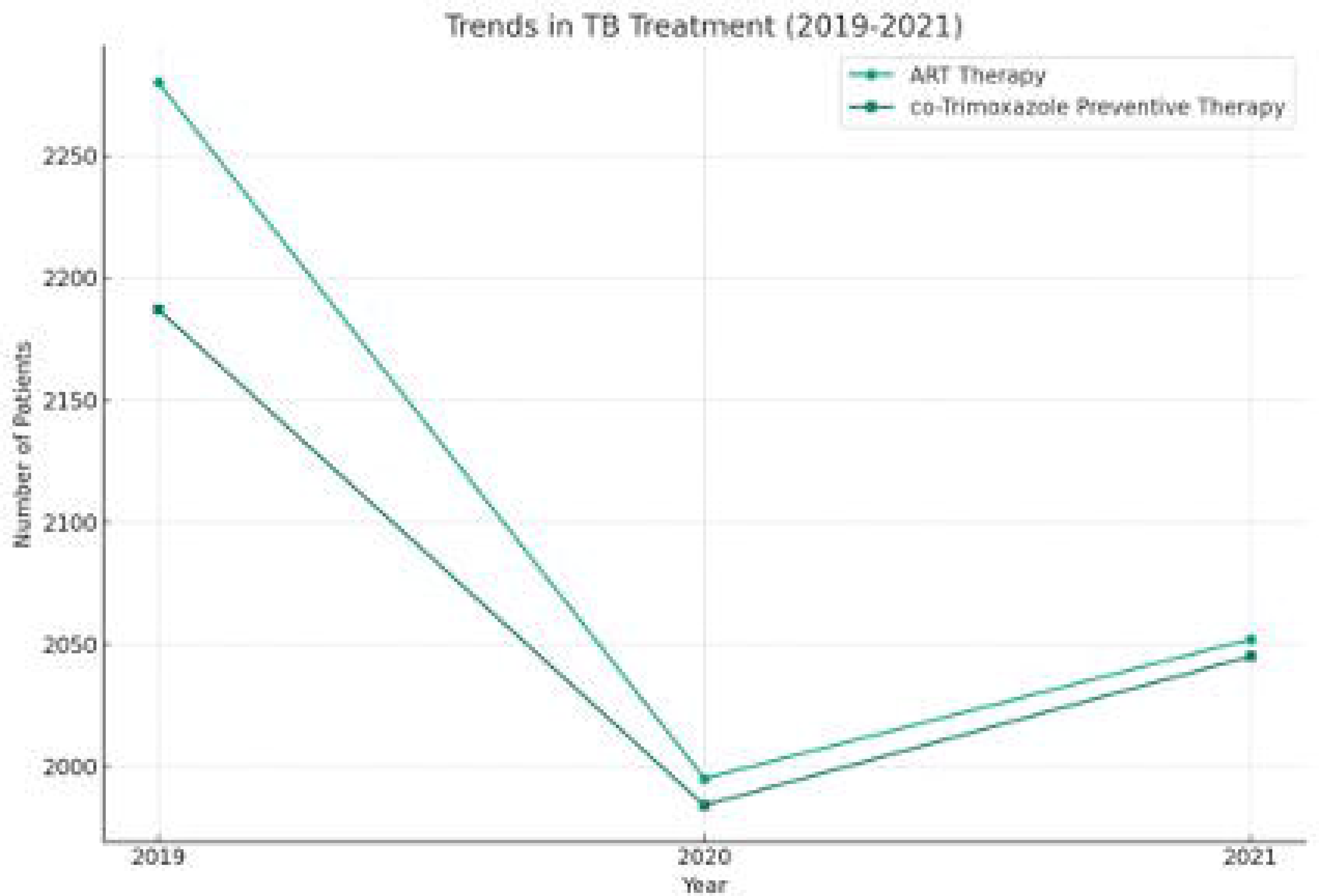
Trends in TB Treatment (2019-2021)

There was a slight increase in the number of patients receiving both forms of treatment from 2020 to 2021. This suggests a recovery or adaptation of TB services, possibly through the implementation of new strategies to deliver care amidst pandemic constraints.

### Inpatient Service Disruption

There was a dive in hospital admissions between 2019 and 2020 when SARS-CoV-2 started in Sierra Leone and a resurgence thereafter (Fig 6). The dive could be attributed to initial shocks from the outbreak.

**Fig.6:**
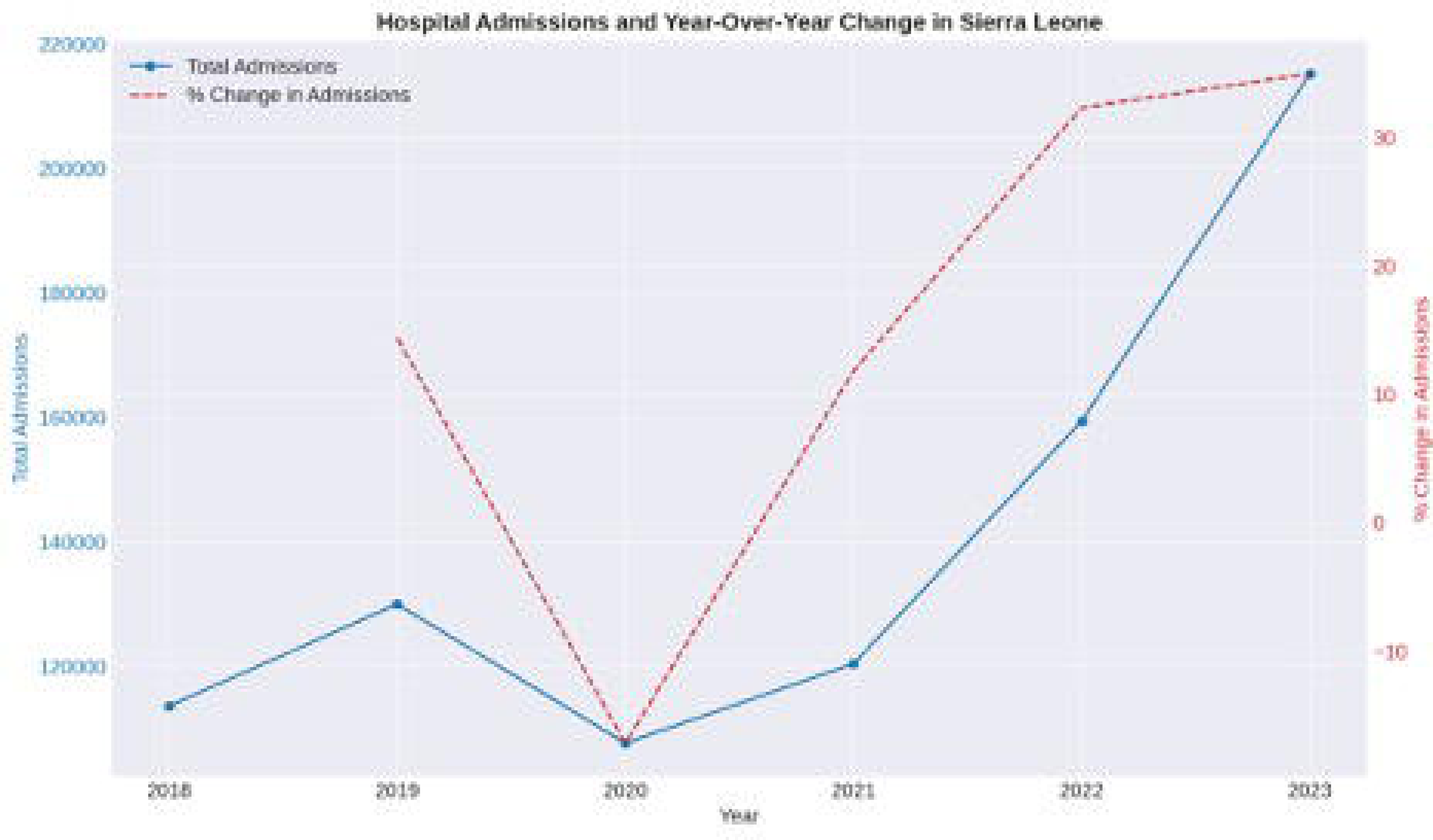
Hospital Admissions and Year-Over-Year Changes in Sierra Leone.

There is likely skepticism at the onset of outbreaks. A focus group discussion in Liberia noted thus: “Whenever they come those who are in front of it, when they tell us, we always have the belief that these people, that money they looking for again so that the notion we always carry. Most of the time we can say that the government naa bring the sickness to put it on the people so we can’t listen to any advice.” This reflects skepticism towards health interventions and a belief that disease outbreaks might be financially motivated or artificially induced, which can significantly affect healthcare seeking behavior and access.

There was a subsequent gradual, substantial rise in healthcare demand during the COVID-19 period, further emphasizing the pandemic’s profound effect on hospital admissions.

There was a mix of increases and decreases in inpatient admissions across different categories from 2018 to 2022. The years 2020 and 2021, which align with the height of the COVID-19 pandemic, showed pronounced changes in admissions, with some categories experiencing significant disruptions (Fig. 7).

**Fig.7:**
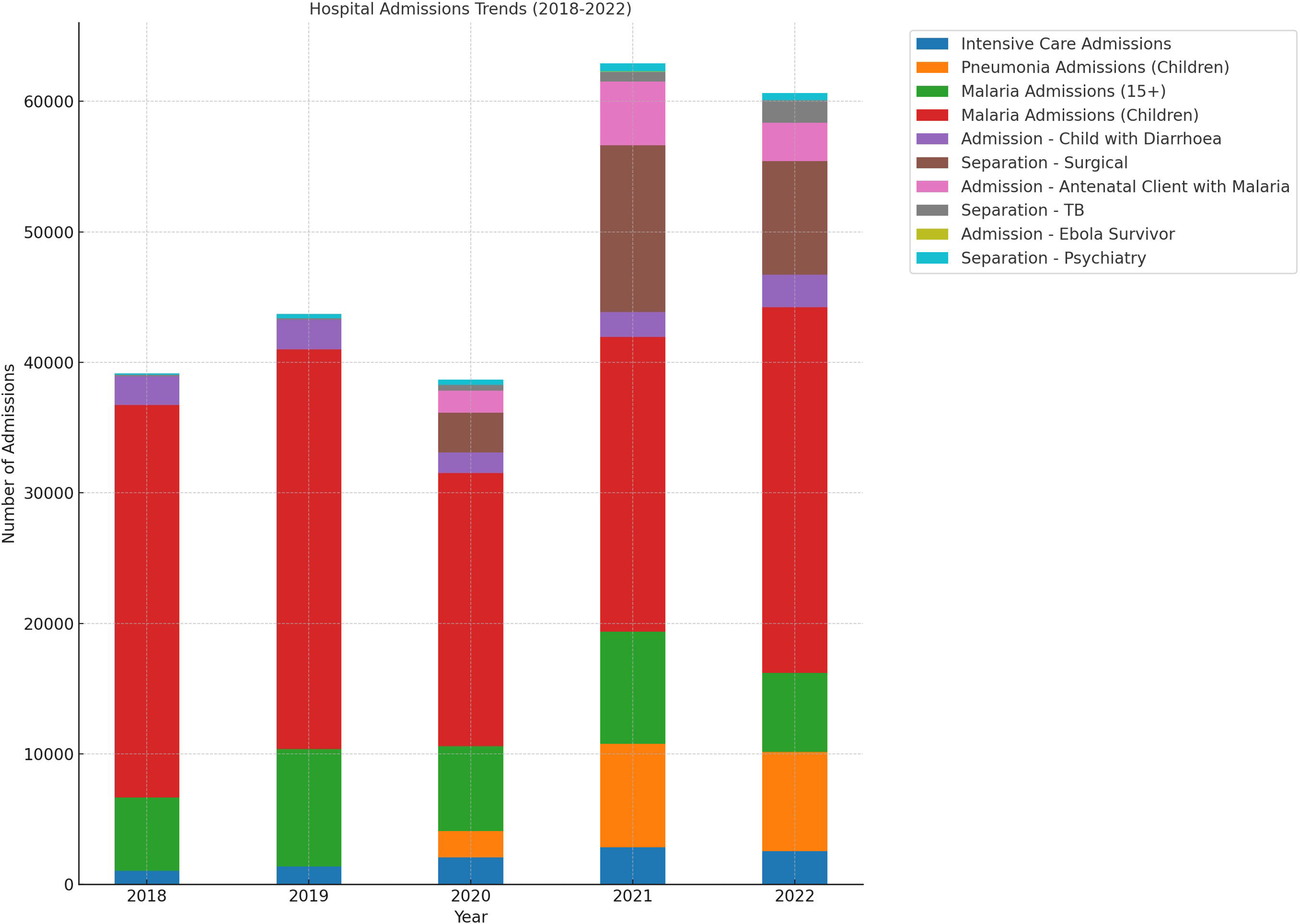
Hospital Admissions Trends.

For the Intensive Care Admissions, a noticeable increase was observed from 2019 to 2021, possibly due to the severity of COVID-19 cases. However, there was a decline in 2022.

There was a significant increase in the number of children with pneumonia admissions in 2021 and a slight decrease in 2022. The symptoms of pneumonia and COVID-19 can overlap, potentially leading to more admissions.

The number of admissions for malaria in individuals aged 15 and above peaked in 2019, declined in 2020 and 2021, and slightly rebounded in 2022. In the case of children with malaria, a decline from 2019 to 2020 was observed, followed by a steady increase in the subsequent years.

The shifts in inpatient admissions could be attributed to multiple factors, including the direct and indirect effects of the COVID-19 pandemic, changes in healthcare-seeking behaviors, hospital resource allocations, and other external events. The increase in intensive care and pneumonia-related admissions during the pandemic underscores the strain on healthcare systems and the potential overlap in symptoms between COVID-19 and other illnesses. Fluctuations in malaria admissions might reflect changes in healthcare access, seasonal patterns, or broader public health interventions.

### Community Perceptions of Healthcare Access and Utilization

The study in West Africa revealed a multifaceted understanding of disease outbreaks, availability of healthcare infrastructure and health practices among community members. Respondents highlighted key insights:

In the case of healthcare access and utilization: challenges in accessing quality healthcare were prominent in all four countries, with many relying on pharmacies or traditional healers due to distant or inadequate health facilities. There was a notable reliance on traditional medicine for ailments during outbreaks.

“Most times, we go to the baba (traditional healer) because the hospital people don’t treat us well….” (Nigeria Qualitative Study Transcription Notes).

In Liberia, lights were shed on the decision to seek care and the gender equal opportunity for community awareness campaigns for care. “In time pass it was just men but right after the Ebola outbreak there are people train in the community to carry on awareness. In case of anything they suspect they create awareness and to advise for the person to go to the facility even though it can be difficult for some of them to go to the facility.” (Liberian In-depth Interview). This reflects a change in who takes charge of health safety in the community, shifting from a male-dominated decision-making process to a more inclusive approach post-Ebola.

Despite the potential gender equal awareness campaigns, at the onset of outbreaks, there is a tendency to boycott the healthcare facilities. “For us when they talk it, we can’t believe it…They will be explaining it to us we be going different way but when it attacks us naa then we will run to the health center.” (Liberian In-depth Interview). This quote suggests a common initial denial of disease outbreaks among community members, followed by a reactive approach to seeking healthcare once the disease has already affected them. Similar observations were noted in Guinea, Sierra Leone and Nigeria.

## DISCUSSIONS

This study has investigated the profound impacts of infectious disease outbreaks on livelihoods, healthcare access, and health outcomes in West Africa, focusing on emerging diseases like COVID-19. Our findings reveal the intricate link between health and socio-economic stability, demonstrating how such outbreaks strain healthcare systems and disrupt socio-economic structures(WHO, 2016).

The recurring outbreaks in West Africa challenge the region’s fragile healthcare infrastructure. Health facilities often face closures or reduced operations due to transmission fears, lack of protective equipment, and workforce attrition, leading to decreased healthcare services (Kieny et al., 2014). This exacerbates the burden of endemic diseases and hampers essential health services like maternal and child care, increasing morbidity and mortality from non-outbreak-related causes (Elston *et al*., 2017).

Moreover, the socio-economic impact of these outbreaks extends beyond immediate health concerns. Livelihoods are severely affected, with communities facing job losses, economic instability, and increased poverty. This disruption in economic activities complicates recovery and resilience(UNDP, 2020).

Our study underscores the need for robust healthcare systems and comprehensive preparedness and response strategies that address both health and socio-economic aspects. Strengthening healthcare infrastructure, ensuring an adequate supply of protective equipment, and maintaining essential health services during outbreaks are crucial(WHO, 2015). Additionally, socio-economic support for affected communities and measures to sustain livelihoods are vital in mitigating the overall impact of such crises(Maxmen, 2020).

## CONCLUSIONS

In conclusion, our research highlights the extensive disruptions caused by infectious disease outbreaks in West Africa, affecting healthcare access, health outcomes, and socio-economic contexts. The recurrent nature of these outbreaks necessitates an urgent and holistic approach to healthcare system strengthening and preparedness(Kieny and Dovlo, 2015).

Our findings call for integrated strategies that address both the health and socio-economic dimensions of outbreak response. Ensuring robust healthcare systems, alongside socio-economic support mechanisms, is essential for the resilience of West African countries(Kieny and Dovlo, 2015; Gebremeskel *et al*., 2021).

Future research should focus on developing and evaluating interventions that effectively bridge the gap between health and socio-economic needs during outbreaks. The lessons learned from West Africa are crucial for global strategies in managing infectious disease outbreaks, emphasizing a multidimensional approach that considers both health and livelihoods(UNDP, 2020).

The resilience of West Africa against future health crises depends on learning from past experiences and preparing proactively for multifaceted challenges. This study contributes to that understanding and advocates for continued efforts to strengthen the region against the challenges posed by infectious disease outbreaks(Maxmen, 2020).

## Data Availability

All data produced in the present study are available upon reasonable request to the authors

## AUTHOR CONTRIBUTIONS

RA, ASB, SC, JG, NM, SM, NH, OD, GM were involved in study conception. RA, ASB, JML, MK were involved in drafting the manuscript. All authors reviewed and revised the manuscript. JML and ASB developed data collection tool. RA, ASB, MK, JML, JG, SC, EC, NM, supervised data collection. RA, ASB, analyzed data.

## ACKNOWLEDGMENTS

We acknowledge all data collectors in Sierra Leone, Guinea, Liberia and Nigeria.

## DATA AVAILABILITY STATEMENT

Datasets generated for this study can be found at https://ona.io/waohecostudy/197244 and https://sl.dhis2.org/. Transcripts of qualitative data are available on request.

